# Circulating Extracellular Vesicles in Human Cardiorenal Syndrome Promote Renal Injury

**DOI:** 10.1101/2023.02.07.23285599

**Authors:** Emeli Chatterjee, Rodosthenis S. Rodosthenous, Ville Kujala, Katia Karalis, Michail Spanos, Helge Immo Lehmann, Getulio Pereira de Oliveira, Mingjian Shi, Tyne W Miller-Fleming, Guoping Li, Priyanka Gokulnath, Ionita Calin Ghiran, JoAnn Lindenfeld, Jonathan D Mosley, Quanhu Sheng, Ravi Shah, Saumya Das

## Abstract

**Background:** Cardiorenal syndrome (CRS)—renal injury during heart failure (HF)—is linked to higher morbidity. Whether circulating extracellular vesicles (EVs) and their RNA cargo directly impact its pathogenesis remains unclear.

**Methods:** Using a microfluidic kidney chip model (KC), we investigated transcriptional effects of circulating EVs from patients with CRS on renal epithelial/endothelial cells. We used small RNA-seq on circulating EVs and regression to prioritize subsets of EV miRNAs associated with serum creatinine, a biomarker of renal function. *In silico* pathway analysis, human genetics, and interrogation of expression of miRNA target genes in the KC model and in a separate cohort of individuals post-renal transplant with microarray-based gene expression was performed for validation.

**Results:** Renal epithelial and endothelial cells in the KC model exhibited uptake of EVs. EVs from patients with CRS led to higher expression of renal injury markers (*IL18*, *NGAL*, *KIM1*) a greater cystatin C secretion relative to non-CRS EVs. Small RNA-seq and regression identified 15 miRNAs related to creatinine, targeting 1143 gene targets specifying pathways relevant to renal injury, including TGF-b and AMPK signaling. We observed directionally consistent changes in expression of TGF-b pathway members (BMP6, FST, TIMP3) in KC model exposed to CRS EVs, as well as in renal tissue after transplant rejection. Mendelian randomization suggested a role for FST in renal function.

**Conclusion:** EVs from patients with CRS directly elicit adverse transcriptional and phenotypic responses in a KC model by regulating biologically relevant pathways, suggesting a novel role for EVs in CRS.

**Trial Registration:** ClinicalTrials.gov NCT 03345446.

**Funding:** AHA (SFRN16SFRN31280008), NHLBI (1R35HL150807-01) and NCATS (UH3 TR002878).

## Introduction

Kidney function is critical to cardiovascular homeostasis. The ability of the kidneys to respond to physiologic and pharmacologic inputs during states of fluid excess or decreased perfusion (e.g., HF) is critical to symptomatic relief and prognosis (1). Although renal dysfunction in patients with HF (CRS) is associated with adverse outcomes (2, 3), therapies against renal hemodynamics [e.g., adenosine receptor agonists (4) or ultrafiltration for fluid unloading (5, 6)] have limited benefit (5). Moreover, renal dysfunction is a risk factor for and develops during therapy in HFpEF and reduced ejection fraction (HFrEF), suggesting the importance of renal reserve across the hemodynamic spectrum. Indeed, despite a mechanistic and therapeutic focus on HFrEF, CRS in HFpEF is equally prevalent, adverse (7, 8), and remains poorly understood. Recent advances in other disease conditions [e.g., cancer (9), diabetes (10)] suggest that trans-organ signaling via circulating extracellular vesicles (EVs) carrying molecular cargo (RNAs, proteins) may be an important mechanism of pathogenesis of different metabolic diseases. Advanced renal dysfunction is potentially associated with increase in EVs bearing pro-inflammatory cargo (11); however, whether these EVs are causal in worsening renal function is clinically relevant but unknown. If circulating EVs in HF promote renal injury, studying their contents will not only unravel potentially novel pathways of renal dysfunction in HF: it may also open novel therapeutic avenues directed at EV-based cargo to maintain renal function during HF therapy and improve outcomes. Nevertheless, the ability to study functional effects of EVs *in vivo* remains limited, given the inability to directly assess the effect of EVs on the human kidney in a dynamic fashion and limited translatable murine models of human renal disease (12, 13). While kidney organoids obtained from human induced pluripotent stem cells (h-iPSCs) model human genetic renal disease (14), the development/induction of hiPSCs into mature kidney-like organoids with mature structures and vascularity remains challenging.

Here, we utilize a recently described *in vitro* system [“Emulate Kidney-Chip”(15)] that recapitulates key features of the blood-kidney interface (renal endothelial/epithelial cells) to test the hypothesis that EVs from individuals with HF with and without abnormal renal function induce molecular phenotypes of renal injury. We characterized plasma circulating EVs from patients with HFpEF with and without renal dysfunction (see **Figure 1** for overall study design). Using microfluidic perfusion to expose renal proximal tubular epithelial/endothelial cells on the Kidney Chip (KC) to EVs from patients with and without CRS, we examined the molecular phenotypic response in two ways: (1) mRNA expression of canonical markers of renal injury *IL18* (16), *NGAL* (17), *KIM1* (18) and function [cystatin C (19)] and (2) expression of computationally identified mRNA targets of HFpEF enriched EV-microRNAs from two sources—the renal cells on the chip (treated with EVs from HFpEF or controls) and human renal transplant biopsy samples with and without renal injury. Our primary findings implicate human HF-derived circulating EV cargo in transcriptional programs are central to renal injury (e.g., related to TGF-β and downstream pathways important for renal fibrosis) and support the translational impact of this technology as a novel method to dissect renal responses.

**Figure 1.**
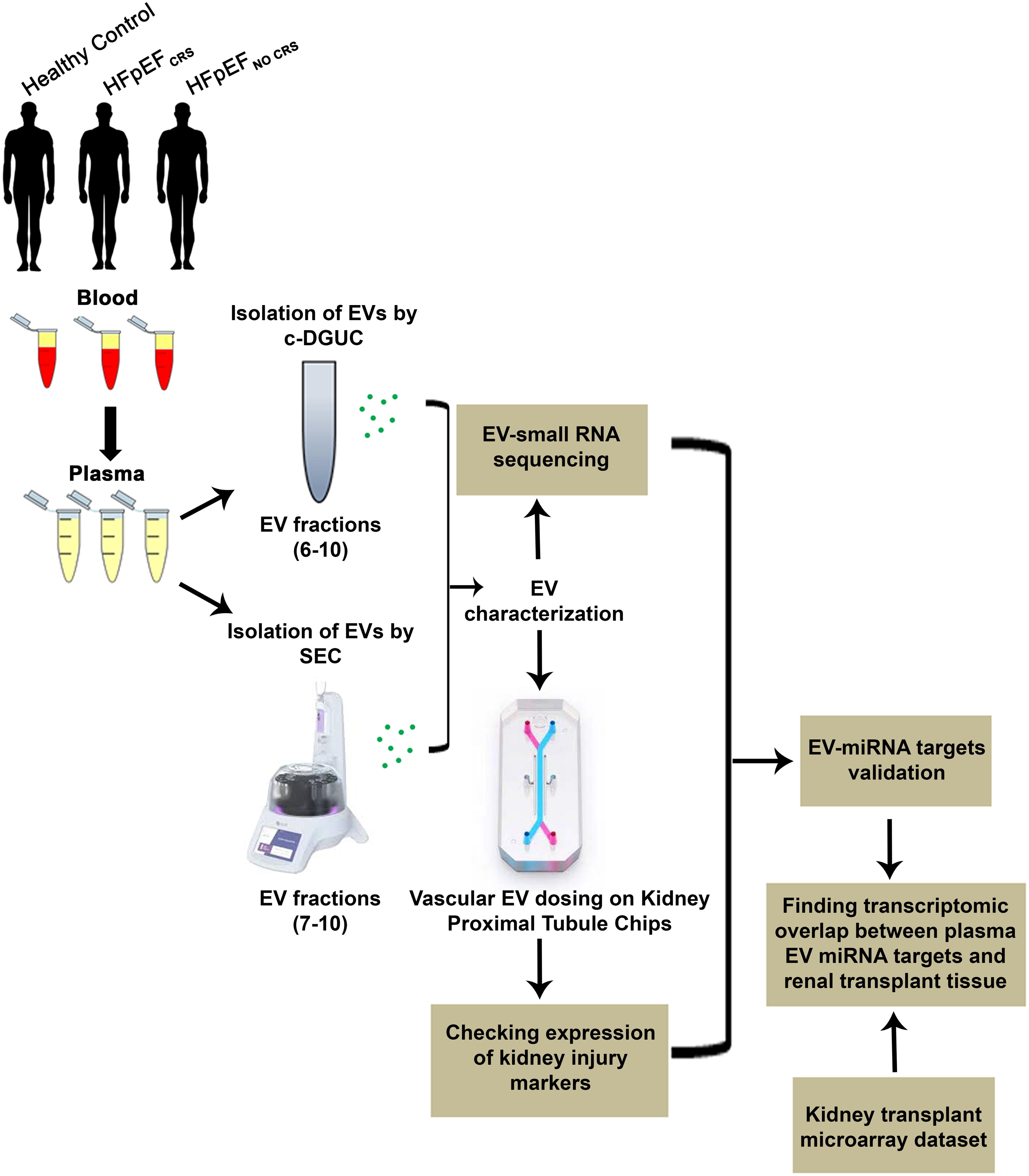
Study schema.

## Results

### Characteristics of study samples

Our study included 12 HFpEF patients and 6 patients without HF (“Healthy Control”) to serve as controls. Of HFpEF patients, 6 met criteria for CRS. Baseline demographic and clinical characteristics are shown in **Table 1** (HFpEF: 83% men with a high occurrence of coronary artery disease, hypertension, and diabetes). Individuals with HFpEF_CRS_ were older (p<0.01) and had increased levels of natriuretic peptides (markers of increased hemodynamic stress) on admission (p=0.01) relative to patients with HFpEF_NO CRS_. In our small RNA sequencing cohort (9 HFpEF; 9 Healthy Control), HFpEF patients were older and had poorer renal function (**Table 2**).

**Table 1.**
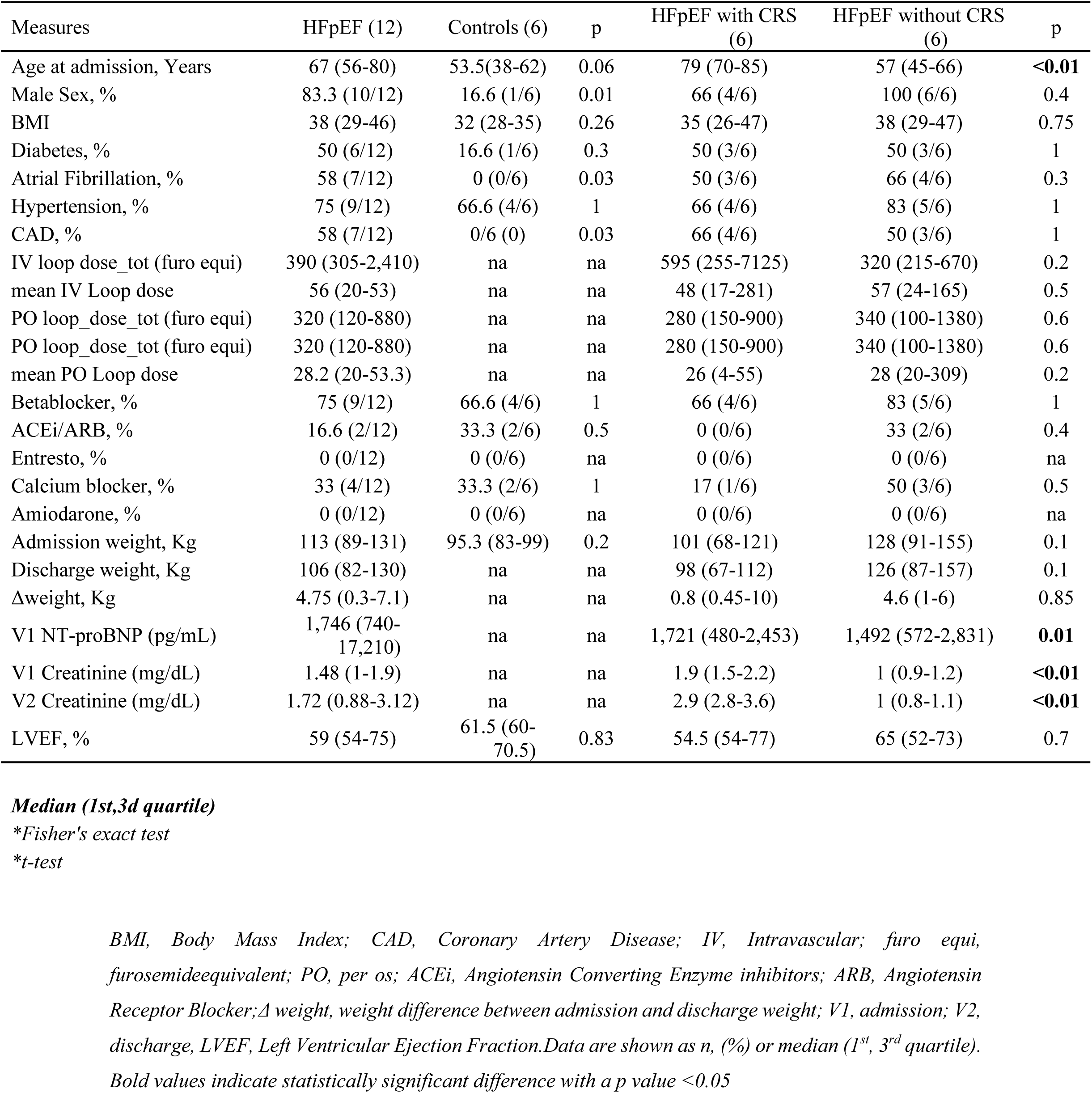
Baseline characteristics of EV treatment cohort- CRS/ non-CRS.

**Table 2.**
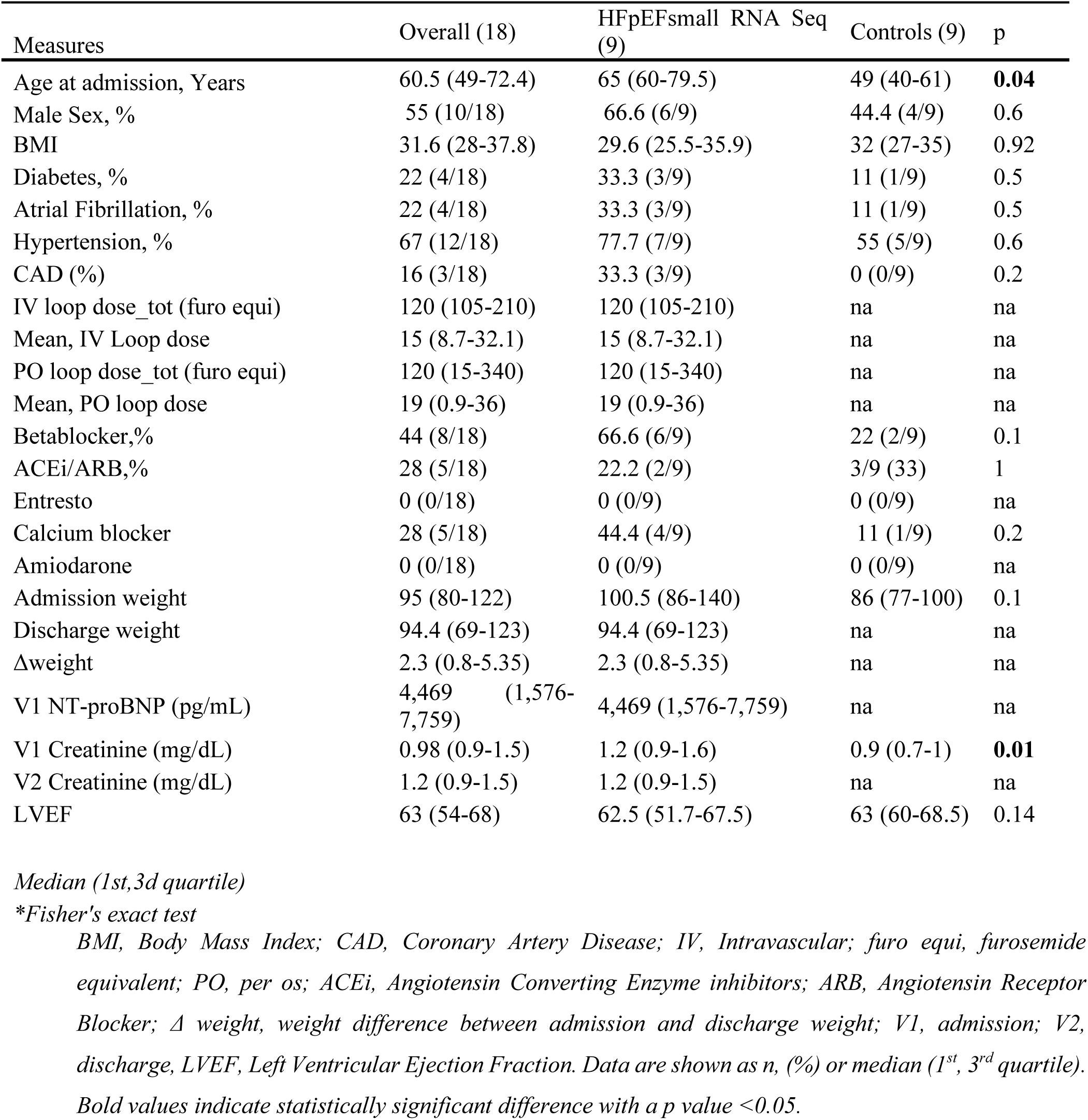
Baseline characteristics of small-RNA Seq cohort.

### Isolation and characterization of circulating EVs

To enhance the rigor of our study, we used two methods [cushion gradient differential ultracentrifugation (c-DGUC) and size-exclusion chromatography (SEC)] for EV isolation from Healthy Control subjects (n=6 for c-DGUC, n=3 for SEC), HFpEF_CRS_ (n=6 for c-DGUC, n=4 for SEC) and HFpEF_NO CRS_ (n=6 for c-DGUC, n=3 for SEC). Isolated EVs were subjected to quality control as specified by MISEV guidelines (20). EV particle number and size distribution were consistent with published morphometric parameters for EVs (particle sizes≈65–100 nm diameter for both c-DGUC and SEC, **Figure 2A**). Canonical EV surface markers (CD63, CD81) and cargo proteins (Alix, Syntenin) were present in pooled EV fractions from both methods, whereas 58K Golgi protein (an indicator of intracellular component contamination)was not found in EVs isolated by either c-DGUC or SEC (**Figure 2B**). Finally, transmission electron microscopy (TEM) confirmed EVs with typical cup-shaped morphology delimited by a double-layered membrane (**Figure 2C**).

**Figure 2.**
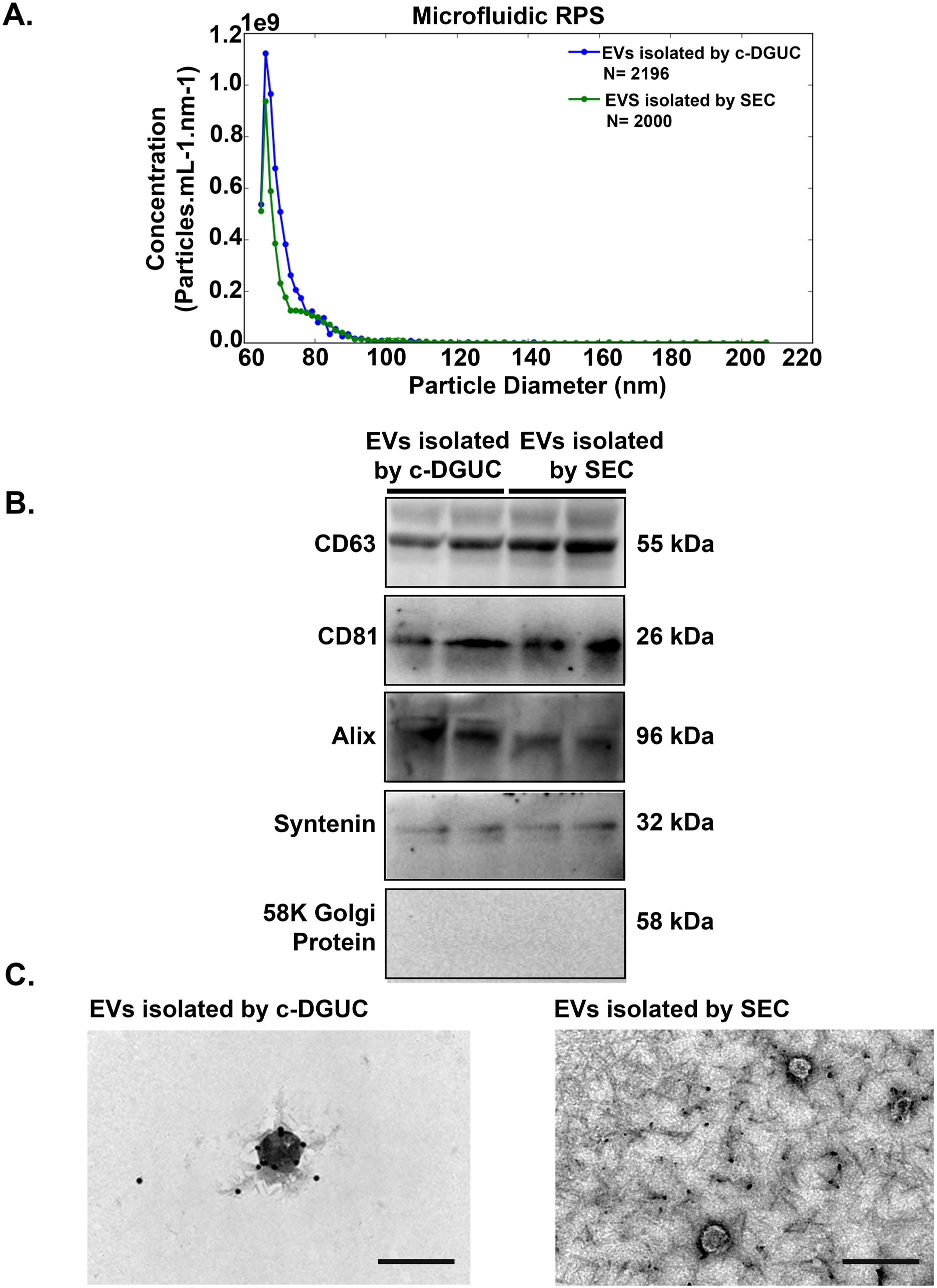
Characterization of extracellular vesicles (EVs) from human plasma: **(A)** Representative microfluidic resistive pulse sensing (MRPS) showing concentration and size distribution profiles of the EV population isolated by c-DGUC and SEC. **(B)**The expression of CD63, CD81, Alix, Syntenin and 58K Golgi protein were determined in the pooled EV samples isolated by both c-DGUC and SEC via western blotting. **(C)** EVs isolated by both c-DGUC **(i)** and SEC **(ii)** were visualized using transmission electron microscope (Scale bar used 200 nm).

### Successful dosing of EVs to *in silico* proximal tubule chip (PCT) model results in differential expression of kidney injury markers

We next studied the effect of isolated EVs on the human Kidney Chip model. The kidney chip utilizes primary human cells to create a physiological model of the human proximal tubule with appropriate interface between epithelial and endothelial cells. Dil-labeled EVs from Healthy Control subjects were observed after 3 days of EV perfusion through the vascular channel via fluorescence microscopy. Fluorescently labeled EVs were abundant within the endothelial (bottom) channel (**Figure 3A**) as well as within the epithelial (top) channel (**Figure 3A**), suggesting uptake of EVs by kidney cells across the chip. **Figure 3B** shows the results of confocal microscopy [**Figure 3B (i)**: endothelial channel; **Figure 3B (ii)**: epithelial channel]. Merged images confirmed abundant uptake of red-labeled EVs into endothelial cells [**Figure 3B (i)**], with lower uptake in the epithelial cells present in the proximal tubule channel [**Figure 3B (ii)**]. These results suggested effective delivery of EVs via microperfusion of the chip.

**Figure 3.**
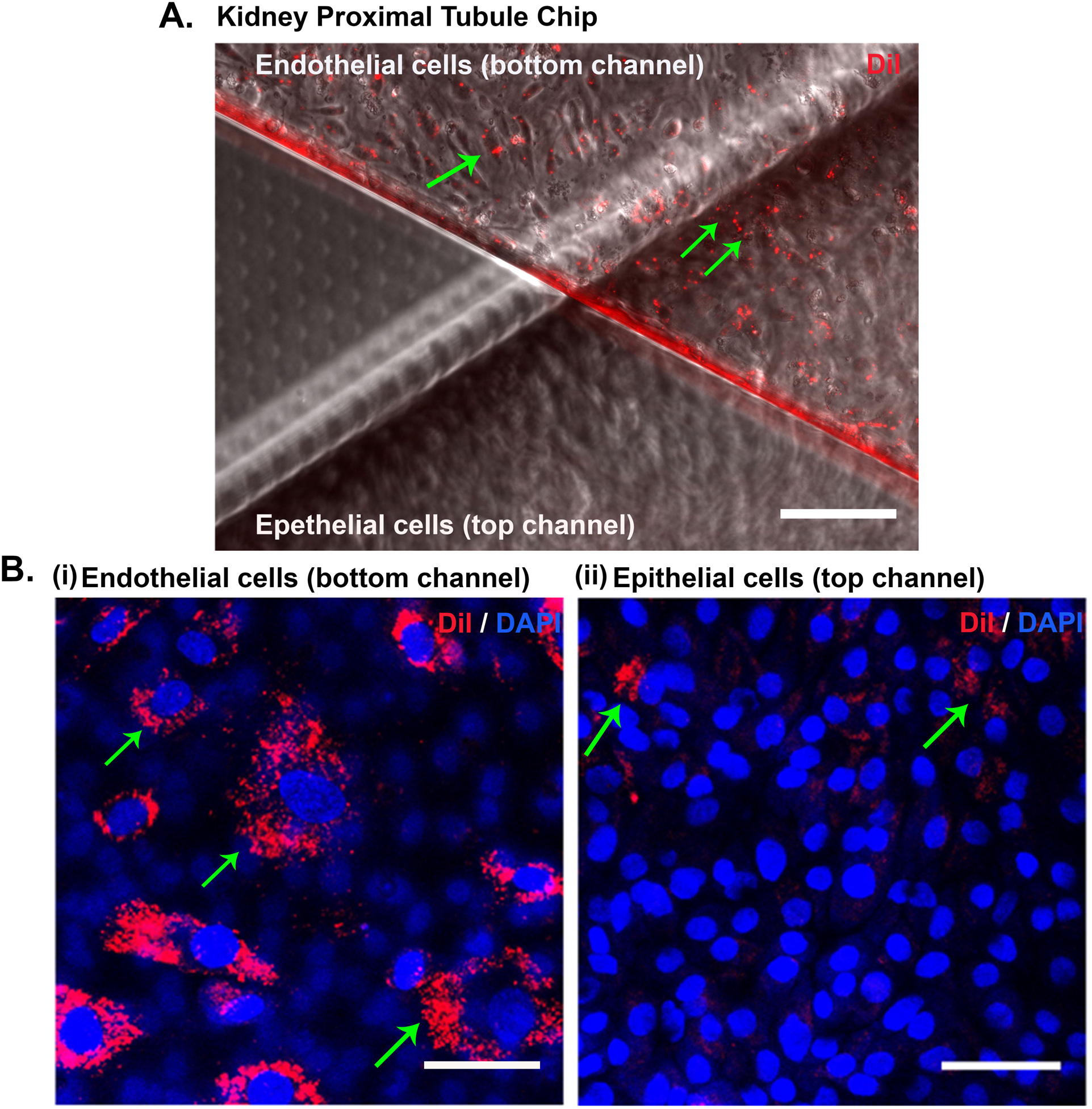
Successful dosing of EVs on Kidney Proximal Tubule Chips: Dil-stained EVs from a Healthy Control subject were visualized after three-day perfusion period using fluorescence microscopy. **(A)** Fluorescently labeled EVs (red), overlaid with a phase contrast image of the chip, can be seen mainly in the vascular endothelial (bottom) channel (Magnification = 100µm). **(B)** EVs were further imaged using a fluorescent confocal microscope. **[B (i)]** Cells in the vascular endothelial channel (bottom) and **[B (ii)]** cells in the epithelial (top) channel were visualized (Magnification = 100 µm).

Next, we sought to determine if EVs from HFpEF patients with CRS functionally affected the cells. We assayed mRNA expression of three canonical markers of renal tubular injury relevant to clinical renal dysfunction: *IL18* (16), *NGAL* (17) and *KIM1*(*18*). *IL18* mRNA expression was significantly higher in endothelial and epithelial renal tubular cells from chips treated with EVs from HFpEF_CRS_ relative to groups either treated with EVs from HFpEF_NO CRS_ or Healthy Control subjects (**Figure 4A, C**). *NGAL* and *KIM1* exhibited similar expression changes with HFpEF_CRS_ EVs (**Figures 4B, D, E**). Results were consistent across the mode of EV isolation (c-DGUC and SEC; **Figures 4A-E**). We assayed protein expression of cystatin C (constitutively expressed across cell types) in effluents coming from the chip model (**Figure 4F**). Changes in cystatin C in the effluent may mimic changes in circulating cystatin C *in vivo* (and reflect changes in renal function) (19). Treatment with EVs from HFpEF_CRS_ significantly upregulated the effluent cystatin C expression of both endothelial and epithelial cells relative to other groups (**Figure 4F**). These results supported an effect of HFpEF_CRS_ EVs on the transcriptional and functional state of the proximal nephron.

**Figure 4.**
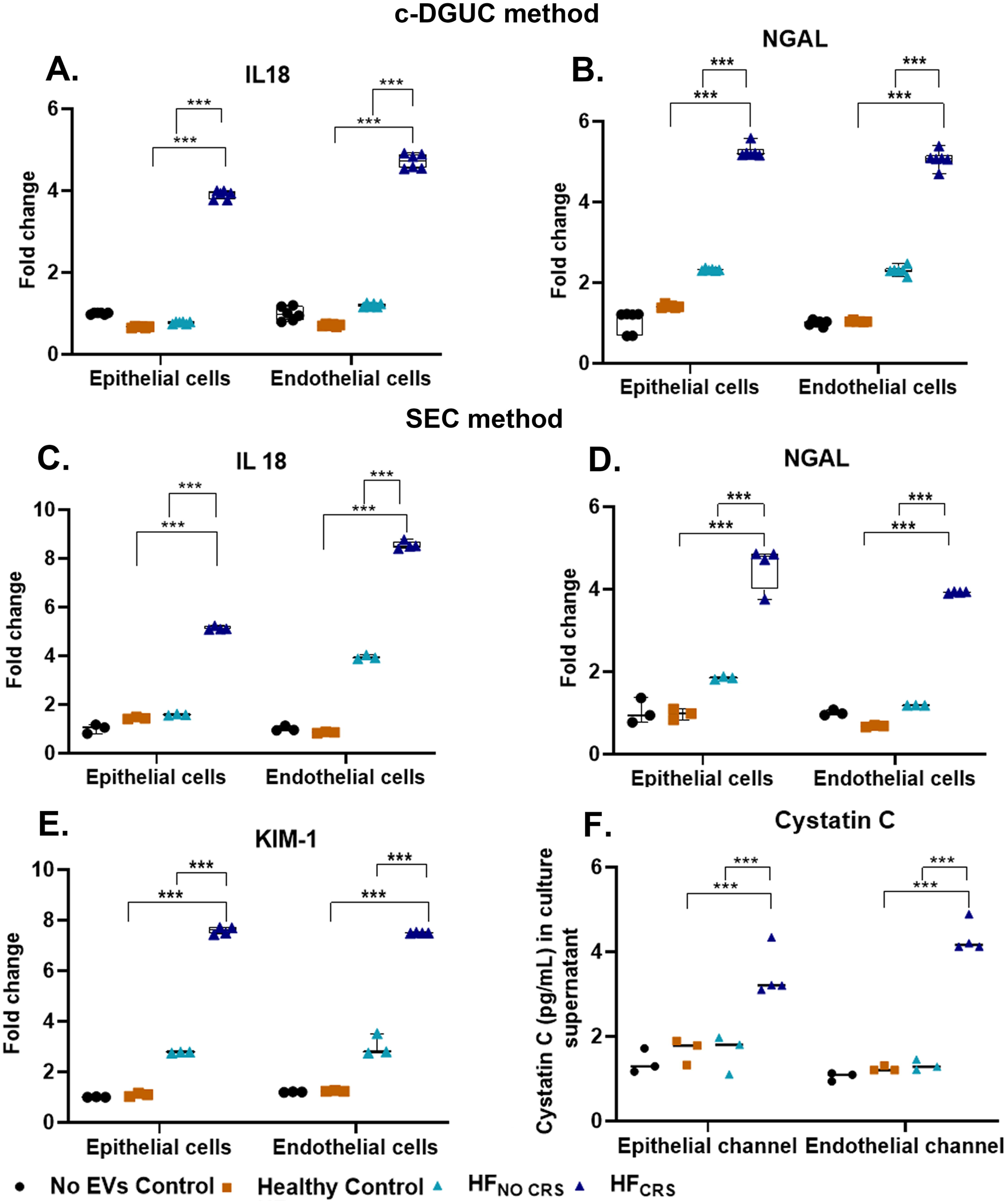
Differential expression of kidney injury marker genes and protein in human glomerular endothelial cells and proximal tubule epithelial cells following 72 hours incubation with EVs. **(A)** Graphical representation of qRT-PCR analysis showing significant higher expression of *NGAL* in both epithelial and endothelial cells of group treated with EVs from HFpEF_CRS_ compared to groups either treated with EVs from HFpEF_NO CRS_ or Healthy Controls. **(B)** Graphical representation of significant higher expression of*IL18* in both epithelial and endothelial cells were observed in group treated with EVs from HFpEF_CRS_ compared to groups either treated with EVs from HFpEF_NO CRS_ or Healthy Control as revealed by qRT-PCR. There is no treatment in “No EVs Control” group. EVs used for the treatment **(A and B)** were isolated by c-DGUC. Three independent chips were prepared for each biological replicates (n=6) of each experimental group. **(C)** mRNA expression of *IL18* was significantly increased in both epithelial and endothelial cells of the group HFpEF_CRS_ compared to groups either treated with EVs from HFpEF_NO CRS_ or Healthy Control. **(D)** The mRNA expression of *NGAL* was significantly higher in the group treated with EVs from HFpEF_CRS_ for both epithelial and endothelial cells when compared with groups either treated with EVs from HFpEF_NO CRS_ or Healthy Control. **(E)** Significant higher mRNA expression of *KIM-1*was evident in both epithelial and endothelial cells of the group HFpEF_CRS_ compared to groups either treated with EVs from HFpEF_NO CRS_ or Healthy Control. **(F)** Graphical representation of cystatin C ELISA showing significant higher expression in the group treated with EVs from HFpEF_CRS_ compared to groups either treated with EVs from HFpEF_NO CRS_ or Healthy Control. EVs used for treatment **(C-F)** were isolated by SEC method. *GAPDH* was used as internal loading control for all experiments. Three independent chips were prepared for each biological replicates (n = 3 for Healthy Control and HFpEF_NO CRS_; n = 4 for HFpEF_CRS_) of each experimental group. Results were analyzed by One-way ANOVA with Tukey’s posthoc test and expressed as ± S.E.M of three independent experiments. ***, p < 0.001.

### Transcriptional targets of heart failure patients derived plasma EVs

MiRNA cargo of EVs can be transferred across many cell types to affect the expression of target genes in recipient cells in many different disease models (21, 22). We studied the extracellular small RNA transcriptome from 9 HFpEF patients and 9 Healthy Controls, demonstrating marked differences in small RNA cargo (more miRNA reads, fewer Y-RNA reads; **Figure 5A**). After mapping reads to the genome (GENCODE GRCh38.p13), 1207 miRNAs were detected. Overall, we observed systematic differences in the miRNA transcriptome of HFpEF EVs relative to that of Healthy Control subjects (**Figure 5B**), with 78 differentially expressed miRNAs detected (at an absolute fold change ≥ 1.5 and 5% FDR) between HFpEF and Healthy Control groups (**Figure 5C**).

**Figure 5.**
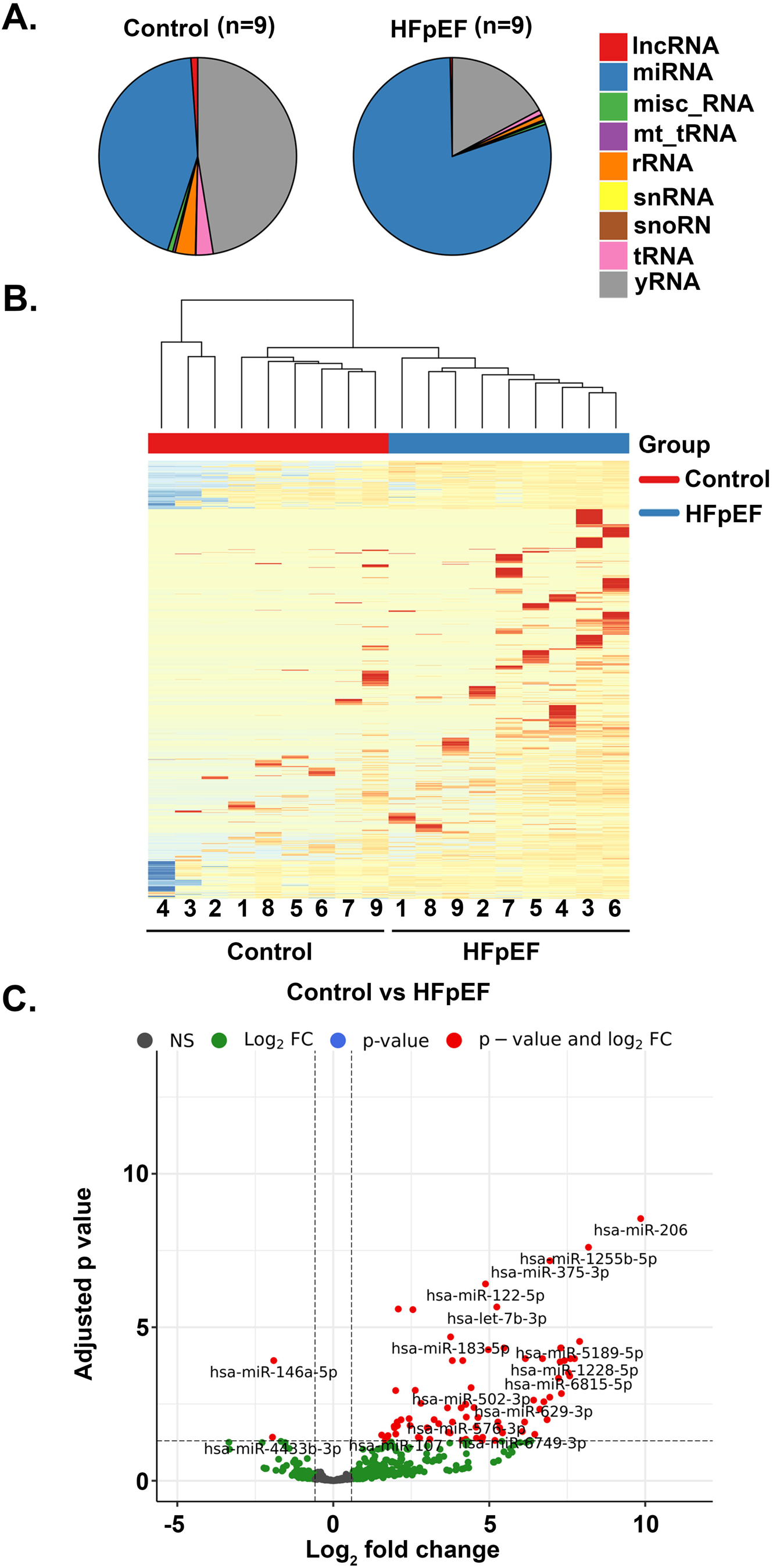
Summary of RNA sequencing results. **(A)** Pie charts showing the differential distribution of ncRNAs according to RNA sequencing in 9 pairs of Healthy Control and HFpEF groups. **(B)** Hierarchical clustering was performed for Controls and HFpEF comparison (n = 9 for each group) based on the differentially expressed genes. The horizontal axis is comprised of all the samples analyzed in the study and vertical axis includes all differentially expressed genes. On Top, control samples are denoted in red squares and HFpEF samples in blue squares. Dark blue to dark red color gradient illustrates lower to higher expression. **(C)** Volcano plot was created by all differentially expressed miRNAs. Y axis shows the adjusted p value and X axis displays the log_2_-fold change value. The red dots represent the differentially expressed miRNAs with FDR adjusted p value ≤ 0.05 and absolute fold change ≥ 1.5, while green dots represent non significantly modulated miRNAs.

### Targeted pathway analyses highlight the TGF-β pathway in human renal dysfunction

We next determined which of the 78 differentially expressed miRNAs within EV were most strongly associated with circulating creatinine levels from all 18 study subjects (a clinical marker of renal dysfunction). We identified 15 out of 78 miRNAs in an elastic net for creatinine (**Table 3**; **Figure 7A, E; Figure S1**). We annotated 1143 high confidence target genes of these 15 miRNAs by multiMiR and performed pathway analysis (DIANA-mirPathV.3) to identify cellular pathways impacted. Thirty-five KEGG biological processes were significantly enriched among putative mRNA targets (**Figure 6**, **Table S2**). Among the different annotated pathways identified, AMPK signaling (23), cell cycle (24), TGF-β signaling (25), and O-glycan biosynthesis (26) were prominent (**Figure 6, Table S2**), supporting a role for perturbation of these signaling pathways central to endothelial-mesenchymal transition/fibrosis in renal dysfunction in HFpEF. Specifically, the “TGF-β signaling pathway” (KEGG) was significantly altered in HF patients with 7 miRNAs (miR-192-5p, miR-122-5p, miR-146a-5p, miR-629-3p, miR-483-3p, miR-378c and miR-21-5p) targeting 27 genes in the pathway. Given its biological relevance in various kidney injury models (25), we prioritized study of the expression of TGF-β signaling pathway genes in the Kidney-Chip cells treated with EVs isolated from Healthy Controls, HFpEF_CRS_ and HFpEF_NO CRS_ via qRT-PCR. *BMP6* (bone morphogenic protein 6), *FST* (Follistatin), *TIMP3* (TIMP metallopeptidase inhibitor 3) mRNA—all targets of miR-192a-5p (<u>higher</u> in EVs from HFpEF; **Figure 7A**)—were <u>down-regulated</u> in both epithelial and endothelial cells of the chip cells treated with HFpEF EVs (**Figure 7B-D**). Conversely, *EGFR* (epidermal growth factor receptor) and *SMAD4* (SMAD family member 4)—targets of miR-146a-5p (<u>down-regulated</u> in HFpEF EVs; **Figure 7E**)—were significantly <u>upregulated</u> in chip cells treated with EVs from the HFpEF patients relative to Healthy Controls (**Figure 7F, G**). While it was not part of the elastic net, we also investigated targets of miR-21-5p, one of the 78 dysregulated miRNAs in the HFpEF EVs, given its role in TGF-β pathway in renal dysfunction (27–29). miR-21-5p was increased in EVs from HFpEF patients (**Figure 8A**), and one of its targets *SMAD7* (SMAD family member 7) was downregulated in both epithelial and endothelial cells after exposure to HFpEF EVs (**Figure 8B**).

**Figure 6:**
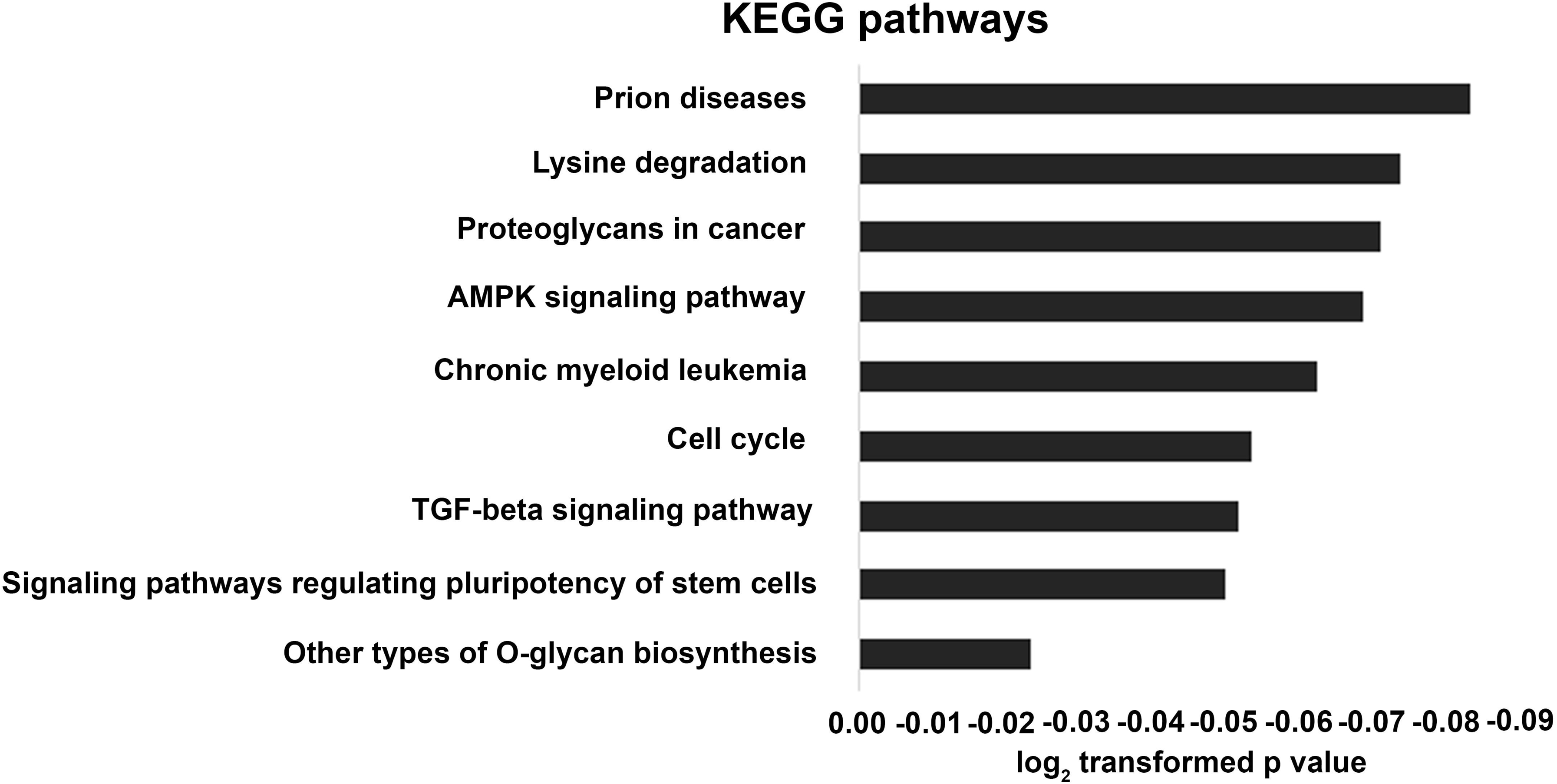
Comparative pathway analysis: **(A)** Bar chart representing 9 most prominent pathways enriched in quantiles with differential EV miRNA patterns in HF compared with Healthy Controls, as revealed by KEGG biological processes.

**Figure 7.**
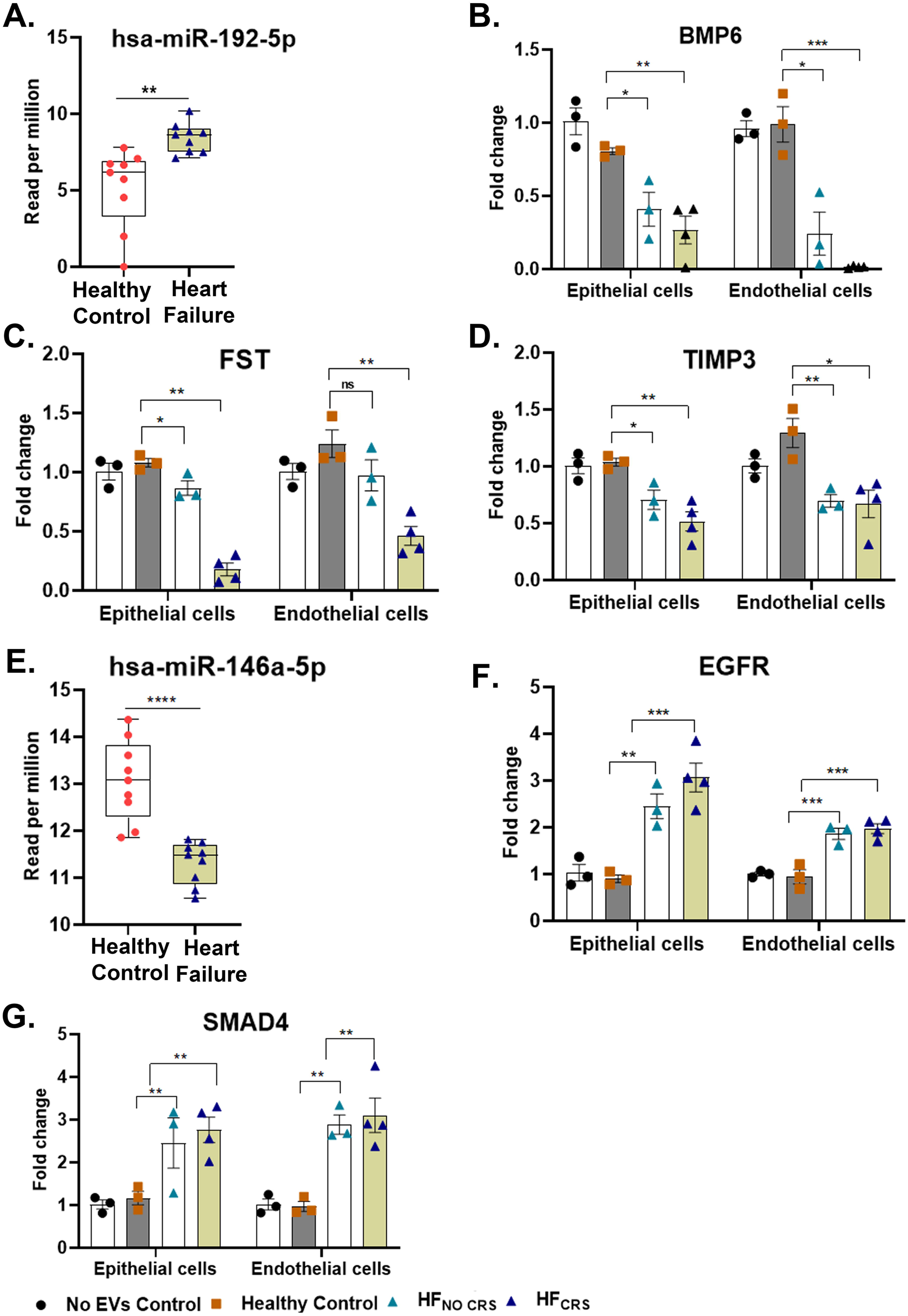
Biological validation of the targets of hsa-miR-192-5p and miR-146a-5p: **(A)** Box and whisker plot showing significant higher expression (reads per million) of hsa-miR-192-5p in Heart failure group compared to the Control group. **(B-D)** qRT-PCR analyses were performed to assay the relative expression profiles of *BMP6, FST* and *TIMP3* genes in Healthy Control and Heart Failure groups. *BMP6, FST* and *TIMP3* were significantly downregulated in either group HFpEF_NO CRS_ or HFpEF_CRS_ compared to Healthy Control group. **(E)** Box and whisker plot showing significant lower expression (reads per million mapped reads) of hsa-miR-146a-5p in HF group compared to the Control group. **(F-G)** *EGFR* and *SMAD4* were significantly upregulated in either group HFpEF_NO CRS_ or HFpEF_CRS_ compared to Healthy Control group. *GAPDH* was used as internal loading control for all experiments. Three independent chips were prepared for each biological replicates (n = 3 for Healthy Control and HFpEF_NO CRS_; n = 4 for HFpEF_CRS_) of each experimental group. Results were analyzed by unpaired *t* test and one-way ANOVA with Tukey’s posthoc test and expressed as ± S.E. of three independent experiments. *, p < 0.05; **, p < 0.01; ***, p < 0.001.

**Figure 8.**
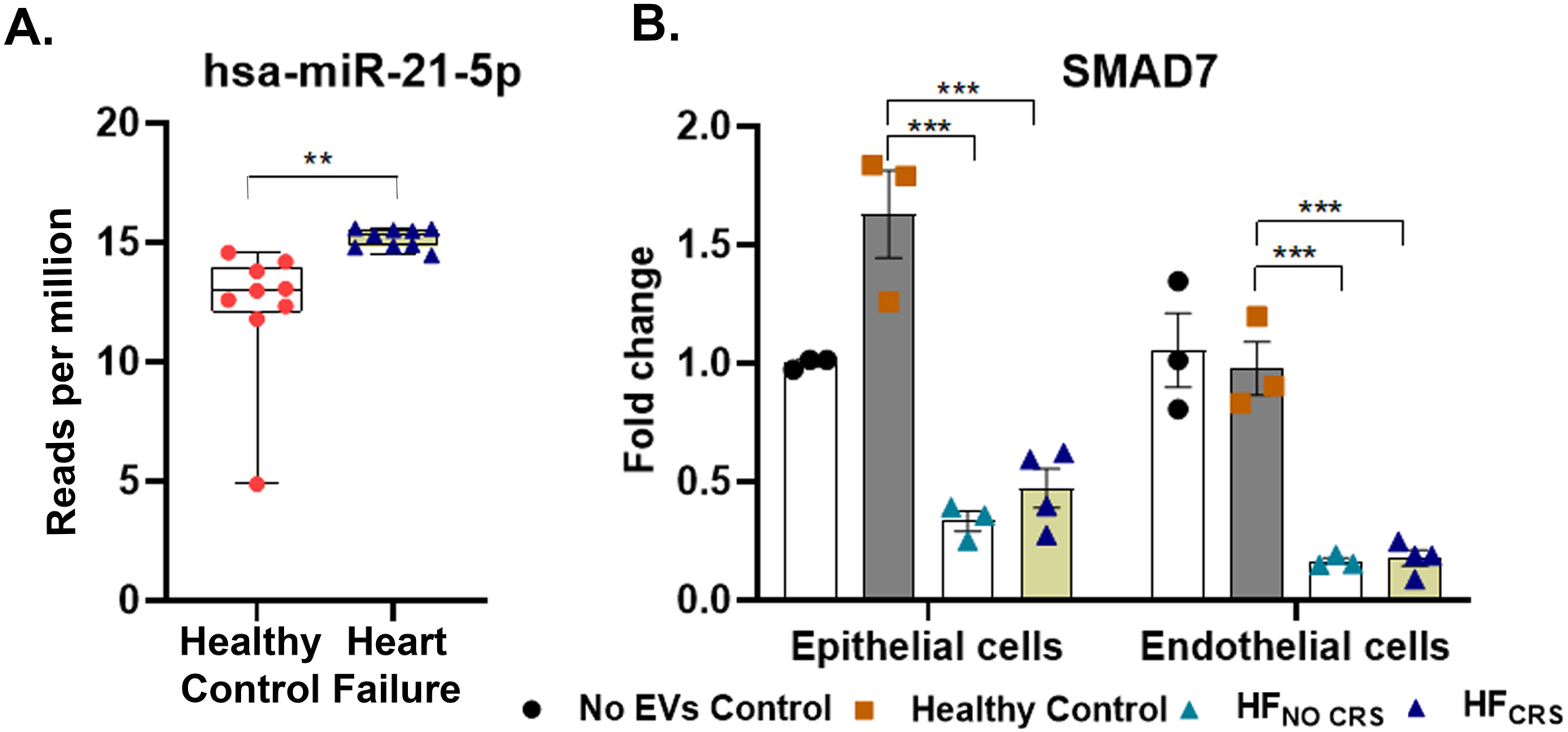
Biological validation of the target of hsa-miR-21-5p: **(A)** Box and whisker plot showing significant higher expression (reads per million) of hsa-miR-21-5p in Heart failure group compared to the Healthy Control group. **(B)** qRT-PCR analysis was performed to assay the relative expression profiles of *SMAD7*gene in Healthy Control and Heart Failure groups. *SMAD7* was significantly downregulated in either group HFpEF_NO CRS_ or HFpEF_CRS_ compared to Healthy Control group. *GAPDH* was used as internal loading control. Three independent chips were prepared for each biological replicates (n = 3 for Healthy Control and HFpEF_NO CRS_; n = 4 for HFpEF_CRS_) of each experimental group. Results were analyzed by unpaired*t* test and one-way ANOVA with Tukey’s posthoc test and expressed as ± S.E. of three independent experiments. **, p < 0.01; ***, p < 0.001.

**Table 3:**
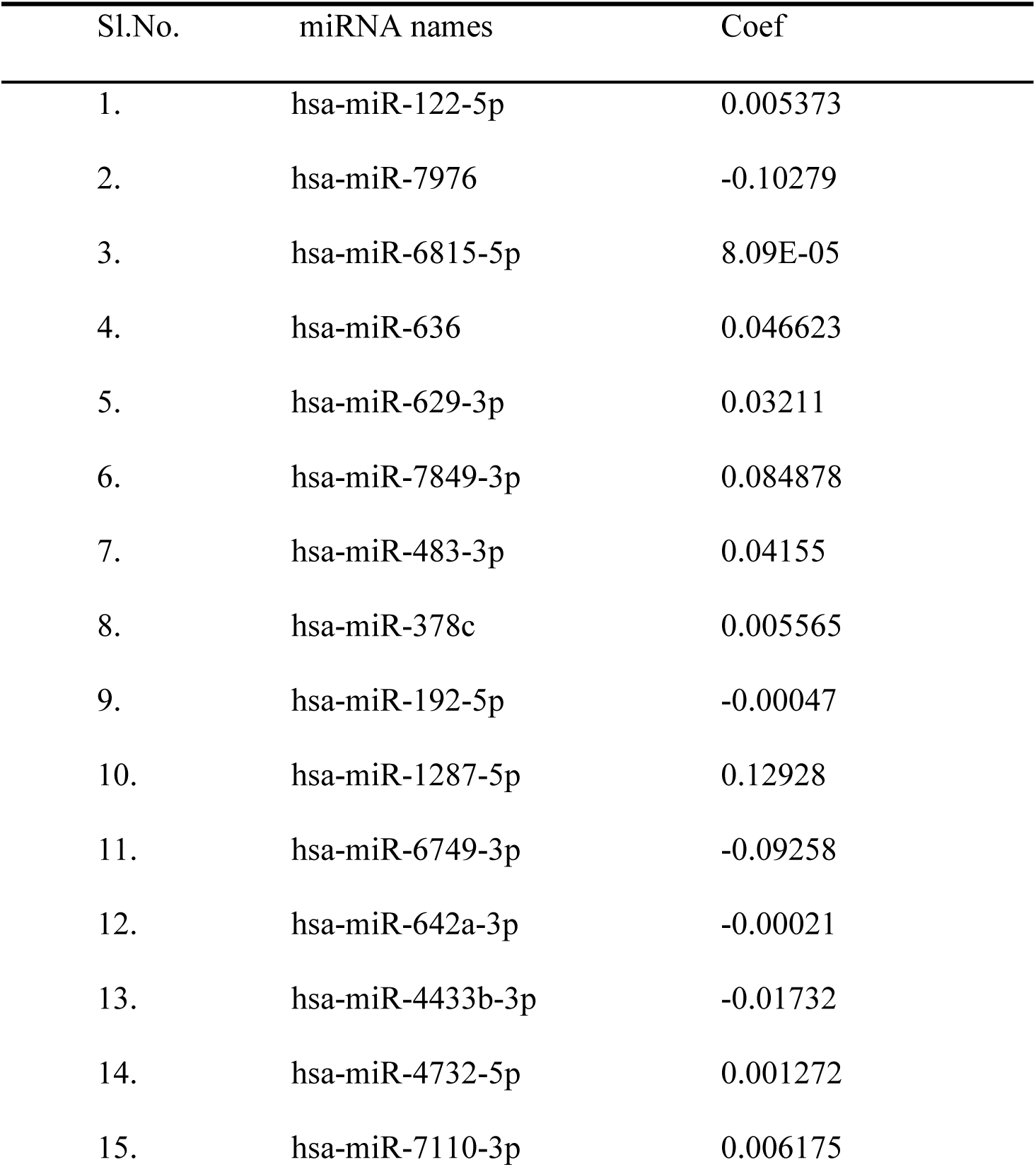
List of miRNAs those were associated with creatinine level.

We next analyzed human tissue in renal transplant patients with or without acute kidney injury undergoing biopsy to identify whether similar cellular pathways identified in our *in silico* model were dysregulated *in vivo* (GEO dataset accession GSE30718). We identified 736 significantly differentially expressed genes between patients with or without kidney injury (at an FDR 5%) out of a total 20848 genes in these samples (**Figure S2A**; enriched KEGG pathways in **Figure S3**, **Table S3**). Interestingly, 1094 out of 1143 mRNAs that were putative targets of the 15 EV-miRNAs associated with plasma creatinine overlapped with the 20848 genes detected in the microarray. Of these 1094 miRNA target genes, 74 genes were present within the 736 gene differentially expressed genes between kidney tissue with or without injury, representing significant enrichment in this dataset (Fisher P < 0.001). Notably, 4 of these genes were related to the TGF-β pathway (**Figure S2B**).

### Expression quantitative trait loci (eQTL) result

Mendelian randomization was used to determine whether genetically determined levels of mRNA expression of the candidate genes were associated with kidney function. There were 2 genes with 1 or more eQTLs that were significantly associated with estimated glomerular filtration rate (eGFR): FST and SMAD7 (**Table S4**). FST demonstrated a significant correlation with eGFR (0.005 [95% CI: 0.002-0.007] mL/min/1.73m^2^ increase in eGFR per standardized unit change in mRNA expression, P=5.5×10^-5^), after adjusting for multiple testing, consistent with a protective effect of higher FST expression on renal function. This was consistent with our observation of downregulation of FST in the KC treated with HFpEF_CRS_.

## Discussion

Cardiac and renal diseases are influenced by synergistic systemic factors (30) and frequently occur concurrently, with amplified clinical consequences for individuals with both. In this context, efforts to resolve how worsening cardiac function influences renal dysfunction are critical to mitigate joint consequences. Studies in CRS have focused on the role of renal hemodynamics and uremia (and accompanying metabolic changes) as a prime driver of renal-cardiac signaling that reinforces myocardial alterations (31), with a broad assessment that overall inflammatory and other uremic toxins during kidney injury lie at the heart of poor cardiac prognosis in HF. This focus on systemic inflammatory and other signaling moieties independent of hemodynamic may be particularly pertinent to HFpEF, where mechanisms of CRS remain poorly understood. While certain shared inflammatory and metabolic stimuli can elicit both renal and cardiac dysfunction in CRS, the potential bidirectional nature of EV interactions (e.g., EVs-renal, and EVs-cardiac signaling) has been less well studied.

In this study, we leverage the use of a human Kidney-Chip to study the functional role of patient derived EVs in CRS associated with HFpEF. The ability to use human model systems to study patient-derived materials to ultimately derive clinically relevant results that can be translated back to human studies is of particular importance in this study given the known short-comings of previous animal and cell culture models. Our study suggests that EVs from HFpEF patients with CRS are directly injurious to renal epithelial and endothelial cells in the short term and can regulate transcriptional pathways that may drive epithelial-mesenchymal transition and renal fibrosis. Notably, our findings focus on the TGF-β signaling pathway and suggest a broad role for this pathway in renal dysfunction/injury in humans. Moreover, genetic alterations in key members of this pathway may predispose to kidney disease.

The fundamental innovation here is the application of a novel microfluidic technology (Kidney-Chip) as a model of renal proximal tubular physiology to characterize EV-based mechanisms of renal injury in HF that begin at the renal-extrarenal interface (circulation). While the concept of an *in vitro* chip-based system to test renal injury has been previously advanced (32–35), its application to evaluate the importance of trans-organ communication in renal injury via EVs is novel. Using high-throughput EV RNA-seq and curated databases of miRNA-mRNA targets, we identified several mRNA targets of differentially abundant miRNAs (between individuals with and without CRS) that are part of the TGF-β signaling network and demonstrated that these specific mRNAs were altered in renal tubular cells on the Kidney-Chip. The TGF-β pathway has been associated in the development of a wide array of kidney diseases as it plays key role in renal fibrosis and inflammation. Moreover, studies also reported that TGF-β/Smad pathway regulates microRNA-mediated renal injury (36).

In line with this, we were able to observe that miR-192-5p and miR-21-5p expression were enhanced in individuals with HF and CRS, with a corresponding downregulation of key targets previously implicated as protective in renal fibrosis [e.g., miR-192-5p: *BMP6* (37, 38), *TIMP3* (39–41); miR-21-5p: *SMAD7* (42–44)]. Previously, it was noticed that miR-192-5p could promote ischemia/reperfusion induced renal injury in rats (45). Previous reports have also revealed that miR-192-5p directs TGF-β mediated collagen deposition during diabetic renal injury via interacting with SMAD-interacting protein 1 (SIP1). miR-21-5p has been considered to play a variety role in regulation of different kidney diseases like allograft dysfunction (46) and diabetic nephropathy (47).

The perturbation of the mRNA targets of these miRNAs in the pathogenesis of renal disease has been previously demonstrated. Indeed, deletion of BMP6 aggravates renal injury and fibrosis by inducing inflammatory cells in renal proximal tubule cells (37). Also, it was observed that, administration of FST as an antagonist of activin can reduce fibrosis during unilateral ureteral obstruction in a preclinical model (48). In line with this, other studies showed that deletion of TIMP3 leads to increased interstitial fibrosis; higher synthesis and deposition of collagen-I suggesting activation of fibroblasts (40). Absence of TIMP3 results in renal injury in murine models (41). In addition to these studies, SMAD7 is shown to play a protective role against a wide range of renal pathology and deletion of SMAD7 leads to diabetic kidney injury (42). SMAD7 also protects from acute renal injury by releasing tubular epithelial cell cycle arrest at the G1 stage during ischemia/reperfusion-induced renal injury *in vivo* (43). Additionally, disruption of SMAD7 results in ANG II-mediated hypertensive nephropathy (44). Finally, miR-146a-5p, a known negative regulator of the TGF-β pathway (49, 50) was downregulated in plasma of individuals with HF and CRS. Fibrosis enhancing gene targets of miR-146a-5p were increased in the renal tubular cells on a chip (*EGFR*, *SMAD4*). SMAD4 plays a key role in regulating TGF-β induced collagen expression and promotes SMAD3-mediated renal fibrosis (51) while activation of EGFR serves as prognostic biomarker during chronic kidney disease (52).

The ability to simultaneously profile circulating EV cargo and to determine their functional implications in the target organ of interest (the kidney) further establishes EVs and their cargo as relevant functional biomarkers of CRS. Prior work studying EVs as biomarkers and mediators of kidney diseases, spans a breadth of conditions [e.g., glomerulonephritis (53, 54), acute and chronic kidney disease (55–58), post-transplant rejection and homeostasis (59, 60), among others]. A consistent finding across studies has been the utility of specific molecular mediators within urinary or circulating EVs as early biomarkers for kidney diseases. For instance, miR-451-5p in urinary EVs in preclinical model of diabetes-induced renal dysfunction appeared to rise before the appearance of albuminuria, a clinical hallmark of glomerular injury in diabetic nephropathy (61). Subsequent studies in human diabetic nephropathy have demonstrated several miRNAs heterogeneously expressed in urinary EVs of patients with diabetes relative to healthy controls (62), with potential implications on fibrosis mechanisms [e.g., miR-320c (62, 63)]. Indeed, several miRNAs identified in urinary EVs have been implicated in renal fibrosis or inflammation [e.g., miR-29c (64), miR-19b (65)]. Our findings here align with and extend beyond these results by not only identifying EV contents, but also by establishing their functional role in renal injury, fibrosis, and dysfunction.

This study represents an innovative first step toward use of novel *in silico* technology to permit isolation of EV effects on physiology in a unique clinical context (CRS). Nevertheless, there are critical limitations in our approach. It is well-accepted that there are a diverse number of extracellular particles in biofluids, including ribonuclear protein (RNP) complexes, lipoproteins, exomeres and supermeres, and that any particular isolation method for EVs may co-isolate these other particles. In that regard, using two complementary isolation methodologies increases our confidence that EVs may indeed be the functional entity in our study. Secondly, there remains some controversy about the carriers for miRNAs in plasma, including their association with RNPs. EVs used in our studies were treated with RNAse to degrade RNA molecules not protected within EVs; nonetheless, it remains possible that the miRNAs we found associated with renal function may be associated with carriers other than EVs.

Certainly, a deeper transcriptional approach with more diverse cell types (e.g., renovascular cells and pericytes) will be critical to model a complex renal cellular ecosystem and broadly cover potential mechanisms of renal injury (e.g., by use of single nuclear RNA-seq in the more complex chip systems). The cell of origin for circulating EVs is also of interest and not discernable with the current design. To elucidate the definitive EV type and cell of origin would require larger amounts of plasma which were not available in this repository. In addition, individuals with HF and CRS had a poorer renal function at study entry, and whether the circulating EVs from these individuals represent a profile of early kidney injury or propensity to progressive injury with therapy for HF remains open. Finally, whether this can be generalized to all forms of HF (e.g., including HF with reduced ejection fraction) and to other comorbid conditions known to influence circulating EV profiles and renal disease (e.g., diabetes, obesity, hypertension) is an area of active interest.

In summary, we leveraged a human Kidney-Chip to decipher the possible contribution of circulating EVs and their RNA cargos in mediating CRS in patients with HFpEF. Our system demonstrated the injurious effect of these EVs and their contents on renal epithelial and endothelial cells and identified key signaling pathways related to TGF-β that may be targeted by miRNAs contained within these EVs (**Figure 9**). Notably, these data add to previous data from animal models that also implicate these pathways in renal injury and are corroborated by complementary human data that suggest an important role for this pathway in renal disease.

**Figure 9.**
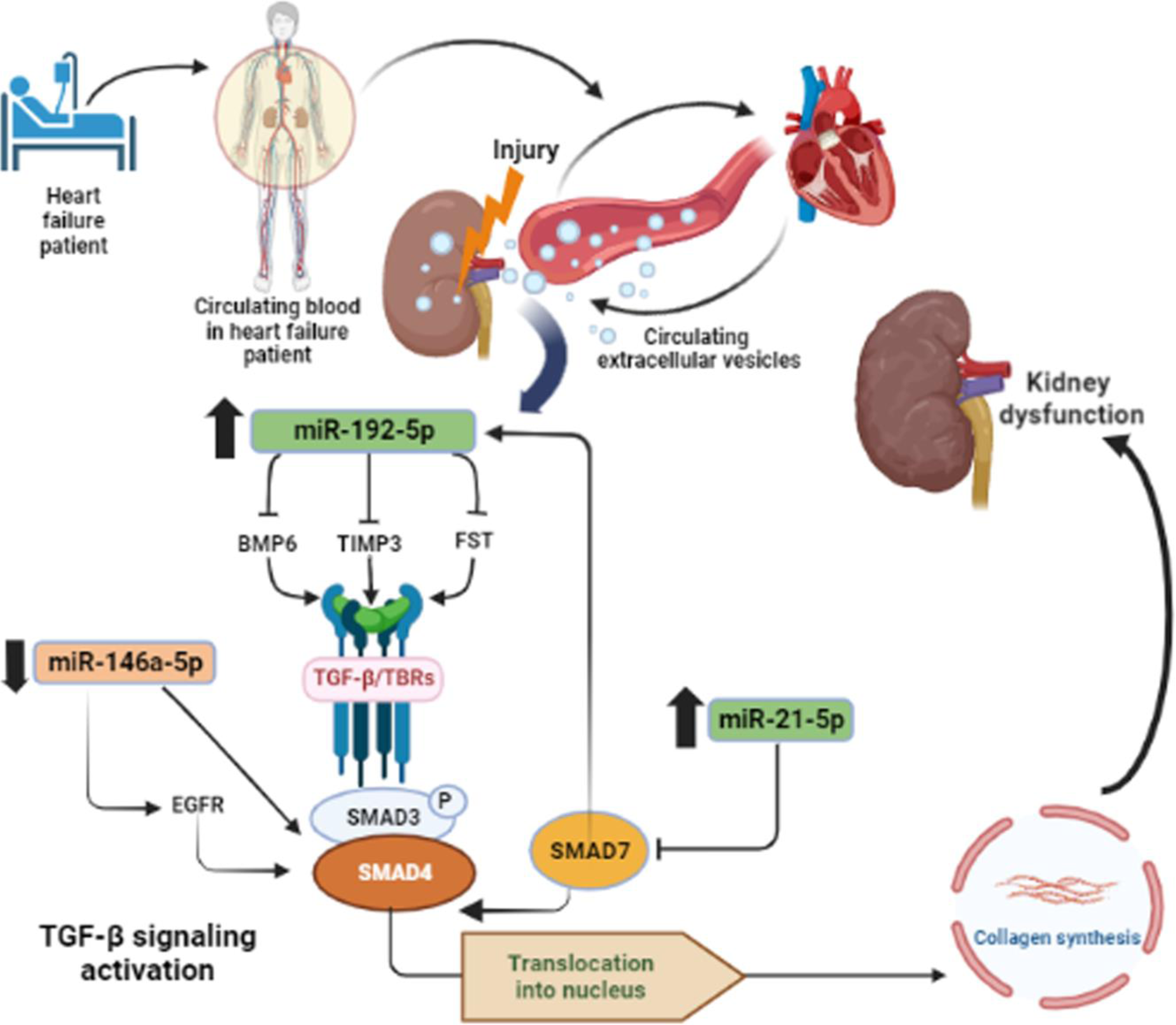
Graphical representation of deleterious effects of plasma EVs promoting kidney injury in human cardiorenal syndrome via targeting TGF-β signaling pathways (created with BioRender.com).

## Methods

### Study population and plasma collection

A total of 12 patients with HF with preserved ejection fraction (HFpEF) with or without CRS were consented under an approved Institutional Review Board (IRB) Protocol (2016P001250), as part of the Circulating RNAs in Acute Heart Failure (Crucial, NCT NCT03345446). HFpEF with CRS [HFpEF_CRS_; 6 patients were used for cushion gradient differential ultracentrifugation (c-DGUC) and out of 6 patients, 4 were again used for size-exclusion chromatography (SEC)] and without CRS [HFpEF_NO CRS_: 6 patients were used for c-DGUC, and out of 6 patients 3 were again used for SEC] were used for the studies. For this study, CRS was defined as present in those individuals with heart failure and renal dysfunction. Following the Acute Dialysis Quality Initiative criteria (7), KDIGO (66) and 7^th^ ADQI Consensus Conference for Definition and Classification of Cardiorenal Syndrome (67), CRS was defined as an increase in creatinine of at least 0.3 mg/dL following admission for acute decompensated HF (Type 1 CRS) or a glomerular filtration rate (GFR) less than 60 mL/min/1.73 m^2^ in the presence of diagnosed HF (Type 2 CRS). Peripheral venous blood was collected at hospital admission and processed within 60 minutes of venipuncture via centrifugation (500g for 10 minutes at room temperature). The supernatant was re-centrifuged at 2,500g for 10 minutes. Plasma samples were stored at −80C until EVs isolation. Plasma from 6 samples for c-DGUC and 3 samples for SEC were included as “control” subjects, as defined by Healthy Controls, were collected following the same protocol.

### Isolation of extracellular vesicles

All plasma samples were processed following either c-DGUC or SEC-based Izon technology (Izon Science, Boston, MA, USA) for EVs isolation, as previously described by our group(21, 68). Fractions 6-10 for c-DGUC and 7-10 for SEC methods were pooled and used for all downstream experiments as described and optimized by our group.

### RNAse A Treatment

Isolated EVs (210 μL) were incubated with RNAse A at a 0.5µg/µL final concentration to degrade any extracellular RNAs not protected within EVs (Thermo Fischer Scientific, Waltham, MA, USA) for 20 minutes at 37 °C followed by an addition of RNAse inhibitor (Thermo Fischer Scientific, Waltham, MA, USA) at a 20U/µL concentration to inactivate RNAse A prior to RNA extraction.

### Microfluidic resistive pulse sensing (MRPS)

EVs were diluted at 1:100 to prevent saturation of the upper limit of detection or aggregation, subjected to MRPS using the Spectradyne’s nCS1 (Spectradyne, Signal Hill, CA, USA), and analyzed with both high and low sensitivity settings (NP100; voltage, 0.60 V; stretch, 46.0 mm and NP400; voltage, 0.40 V; stretch, 43.5 mm respectively). The pressure was preset at 7.0 mbar. Minimum 2000 particles were analyzed for each sample.

### Western blot for EVs isolated through c-DGUC and SEC

Western blot analysis was done as described(21). Concentrated EV suspensions from plasma were lysed for protein extraction (RIPA lysis buffer; 1X protease and phosphatase inhibitor cocktail, Thermo Fisher Scientific, Waltham, MA, USA) for 20 minutes at 4°C. Protein concentration was quantified with Pierce BCA Protein Assay Kit (Thermo Fisher Scientific, Waltham, MA, USA) followed by SDS-PAGE. Gels were transferred to PVDF membranes (Millipore Sigma, Burlington, MA, USA) and blocked with 5% bovine serum albumin (BSA; Millipore Sigma, Burlington, MA, USA) for 1hour at room temperature. Primary antibodies [CD81 (Biolegend, San Diego, CA, USA), CD63 (Abcam, Cambridge, UK), Alix (Abcam), Syntenin (Abcam) and 58K Golgi protein (Abcam)] were incubated at 4°C rocking at a 1:1000 concentration. Secondary HRP-antibodies (Cell Signaling Technology, Danver, MA, USA) were incubated for 1hour at room temperature rocking. Blots were developed using the Super SignalFemto developer (Thermo Fisher Scientific, Waltham, MA, USA).

### TEM of plasma EVs

We performed TEM to define the shape and size of isolated EVs. A drop containing 5 μL of purified plasma EVs (for both c-DGUC and SEC) was placed on parafilm, and a carbon-coated copper grid was placed on top of the drop for 30 minutes. Carbon-coated copper grids were previously glow discharged for 30 seconds to turn into an overall hydrophilic surface. For immune gold labeling, grids were blocked with 1% BSA in 1X PBS for 10 minutes, and IgG primary antibody anti-CD81 (Santa Cruz Biotechnology, Dallas, TX, USA; 1:30 dilution in blocking reagent) was used for 30 minutes at room temperature. Grids were then washed three times in 1X PBS and incubated with Protein A, conjugated with 10 nm gold particle (1:30 dilution) for 20 minutes at room temperature. Grids were washed twice with 1XPBS (for 5 minutes total) and 4 times with water (for 10 minutes total). Grids were stained with 0.75% uranyl formate for 1 minute and visualized on a JEOL 1400 electron microscope outfitted with an Orius SC1000 CCD camera (Gatan, Inc. Pleasanton, CA, USA).

### *In vitro* renal model (Kidney-Chip)

The goal of the “Kidney-Chip” technology is to simulate the microenvironment in the proximal nephron, including exposure of the renal environment to circulating plasma (endothelial interface) and the functional response of the renal epithelium to these contents (epithelial surface). By design, each chip includes epithelial cells in the apical channel and endothelial cells in the basal channel. These two channels are parted by a porous membrane which allows the cell-to-cell interaction mimicking the *in vivo* system.

To construct the human Proximal Tubule Kidney Chip, polydimethylsiloxane (PDMS) chips (Chip-S1; Emulate, Boston, MA, USA) were used. This chip contains two fluidic parallel channels: an upper channel or apical channel with a dimension of 1mm high × 1mm wide and a lower or basal channel with a dimension of 0.2 mm high × 1 mm wide. The pore diameter of the parting membrane is 7.0µm and thickness is 50µm. These two-channel, microfluidic chips were surface activated with ER-1 solution (Emulate) according to the manufacturer’s protocol. Both apical and basal channels were coated with ECM solution [Collagen-IV 0.05mg/mL (Sigma, Burlington, MA, USA) and 0.1mg/mL Matrigel (Corning) in DPBS (Gibco)] overnight at 37°C. On the following day, the bottom and top channels of the chips were filled with Human Renal Microvascular Endothelial Cell (hRMVEC; Cell Systems, Kirkland, WA, USA) and Human Renal Proximal Tubule Epithelial Cell (hRPTEC; Lonza, Basel, Switzerland) media respectively followed by seeding with hRMVECs (2 x 10^6^ cells/mL) to the bottom channels and were incubate at 37°C for 2.5-3 hours in a “flipped” condition to facilitate cell attachment. On the next day, hRPTECs (1.0 x 10^6^ cells/mL) were seeded to the top channel and incubated overnight at 37 °C (confirmed by microscopy). In effect, the upper channel simulated a tubular lumen lined by primary human epithelial cells seeded on an ECM coated membrane, while the basal channel was lined with endothelial cells to simulate peritubular vasculature. A gravity wash was performed to confirm that the nutrients were filled and that the channels did not desiccate. Chips with cells seeded were then cultured under a “degassed” (a process of passing warmed media through a Steriflip^®^ filter unit to remove the dissolved gases from aqueous solutions) media flow (30 µL/hour) rate with no stretch applied (0 Hz, 0%) using an automated culture system (Emulate). Chips were maintained for another 96 hours at this condition before EV experiments.

### Application of human-derived EVs to the Kidney-Chip

In this study, since EVs were isolated from the blood samples, our aim was to mimic the perfusion of the vasculature of blood vessels during EV dosing. Successful EV dosing should result in significantly higher uptake of EVs to the bottom channel compared to top channel.

To determine whether the EV uptake within the chip was successful, purified EVs from Healthy Control subjects were labeled with a tracer dye, Dil (5 mmol/L, Thermo Fisher Scientific, Waltham, MA, USA) for 20 minutes at 37°C. To get rid of the excess dye, EVs were centrifuged at 750g for 2 minutes using a spin column (Exosome Spin Columns, MW 3000, Thermo Fisher Scientific, Waltham, MA, USA) and re-suspended in 1X PBS. This was repeated twice. Dil stained EVs were diluted at 1:50 into degassed complete endothelial media and added as a single bolus to each bottom inlet (endothelial surface) after aspirating all media from both inlets and outlets followed by uninterrupted flow for 3 days. We designated “No EVs Control” chips without exposure to the EVs (but with endothelial media) that were treated identically.

### Effluent sampling from the chips

Effluents were collected from all Pod outlet reservoirs, avoiding direct contact with reservoir “Vias” stored in pre-labeled appropriate multi-well plate, and placed on ice immediately. The amount of cystatin C in the sample effluents of different groups (HFpEF_CRS_, HFPEF_NOCRS_, Healthy Control and No EVs Control) was quantified via ELISA (Abcam, Cambridge, UK) following manufacturers’ protocol (expressed as pg cystatin C/mL cellular effluent).

### RNA extraction and quantification

After 3 days of dosing chips with human-derived EVs, the chips were disconnected, washed with 1X PBS, and filled with RNAlater (Invitrogen, Waltham, MA, USA) to preserve cells for RNA extraction. The PureLink RNA Mini Kit (Thermo Fischer Scientific, Waltham, MA, USA) was used following the manufacturer’s protocol. Total RNA was eluted in 20µL, treated with DNAse, and “cleaned-up” using RNA Clean & Concentrator-5 with DNase I (Zymo Research, Irvine, CA, USA) according to the manufacturer’s protocol. Final RNA concentration was quantified by spectrophotometry (Nanodrop 2000, Thermo Fischer Scientific, Waltham, MA, USA).

The High Capacity cDNA Reverse Transcription Kit (Thermo Fischer Scientific, Waltham, MA, USA) was used for cDNA synthesis from RNA. For amplification and quantification of selected genes (*IL-18*, *BMP6*, *FST*, *TIMP3*, *EGFR*, *SMAD4*, *SMAD7*), the ExiLENTSYBR® Green master mix (Exiqon, Vedbæk, Denmark) was used on a QuantStudio 6 Flex Real-Time PCR System up to 40 amplification cycles. Any amplification cycle (Ct) greater than or equal to 40 was assigned as a “negative threshold” and was therefore not included in our calculations. Relative gene expression was quantified by (2^-ΔΔ^Ct) method after normalization of genes of interest to the internal control *GAPDH*. All qRT-PCR primer sequences are summarized in **Table S2**.

### Small RNA sequencing of plasma EV samples

We performed Small RNA sequencing on the plasma of patients with HFpEF or Healthy Controls to identify differences in extracellular RNA cargo. The methods of RNA-seq have been previously published by our group and are reproduced from there with little editing to allow for scientific replication and rigor, with this statement serving as the attestation (69). Samples from 9 Healthy Control subjects and 9 patients with HFpEF were sequenced, applying the same inclusion criteria for this analysis. Extracellular RNA was isolated from 1 mL of plasma sample and small-RNA libraries were constructed following our previously published methods (69). Plasma exRNA was isolated using the exoRNeasy Serum/Plasma Midi Kit (Invitrogen, Waltham, MA, USA) and libraries were constructed from approximately 10 ng RNA using the NEBNext Small RNA Library Prep Set for Illumina (New England Biolabs, Ipswich, MA, USA). Size selection of libraries was performed by gel electrophoresis on a 10% <u>Novex</u> TBE gel with excision of the 140 to 160 nucleotide bands (corresponding to 21–40 nucleotide RNA fragments). Libraries were diluted to a final concentration of 2 nM, pooled, and sequenced on an Illumina HiSeq 2000 for single read 50 cycles.

Bioinformatic processing was performed using TIGER, as described (70). Briefly, Cutadapt (v2.10) (71) was used to trim 3’ adapters for raw reads. All reads with less than 16 nucleotides were designated as “too short” and discarded. Quality control on both raw reads and adaptor-trimmed reads was performed using FastQC (v0.11.9) (www.bioinformatics.babraham.ac.uk/projects/fastqc). The adaptor-trimmed reads were mapped to the GENCODE GRCh38.p13 genome, addition to rRNA and tRNA reference sequences, by Bowtie (v1.3.0) (72) allowing only one mismatch. Significantly differential expressed miRNAs between HPpEF and Healthy Control samples with absolute fold change ≥ 1.5 and FDR-adjusted p-value ≤ 0.05 were detected by DESeq2 (v1.30.1) (73) using total host smallRNA reads as normalization factor.

### Identification of miRNAs associated with creatinine

We used elastic net regression (linear) to identify a parsimonious model of miRNAs associated with circulating creatinine levels. Log-transformed reads per million expression of the significantly differential expressed miRNAs were included as independent variables in an elastic net regression for creatinine level (in mg/dL) as the dependent variable (*glmnet* in R). The miRNAs included in the final elastic net were used for target gene annotation using the multiMiR package. Only the genes annotated as target genes in at least two out of three databases, including mirecords, mirtarbase and tarbase, were retained as high confidence target genes.

### Human renal transplant biopsy samples

To determine if the mRNA targets of the EV miRNAs associated with creatinine (by elastic net) in HFpEF were deregulated in human kidney tissue, we queried a published microarray data set of renal transplant patients with acute kidney injury [GSE30718 (74)]. From this study, 28 individuals with acute kidney injury (“sample” group) and 11 “pristine” protocol biopsy samples (control group) of the microarray dataset (GSE30718) were analyzed based on GEO2R R script with following modifications: (1) the probes without gene symbol annotation and probes mapped to multiple gene symbols were discarded before differential expression analysis, and (2) for gene symbols mapped by multiple probes, the probe with the smallest p value was kept as the representative. Significantly differentially expressed genes with absolute fold change ≥1.5 and FDR-adjusted p-value ≤ 0.05 were detected by linear model using the limma package. The differentially expressed genes were used in KEGG pathway over-representation analysis (ORA) by WebGestaltR package. Fisher exact test was used to test the enrichment of the differentially expressed genes in miRNA target genes comparing to all microarray genes.

### Expression quantitative trait analysis

SNPs associated with mRNA levels for each of the 6 candidate genes (BMP6, EGFR, FST, SMAD4, SMAD7, TIMP3) were identified using data from the GTEx version 8 resource (75). The best performing gene expression model for each gene was identified by selecting the model with the highest performance r^2^, comparing PrediXcan, UTMOST, and JTI methods for gene expression imputation (76), (77), (78). The SNPs identified by the best-performing model were then used in the downstream eQTL analysis. The association between each of these expression quantitative loci (eQTLs) and kidney function was examined using GWAS summary statistics of eGFR (n=1,201,909) (79), and those genes with 1 or SNPs associated with eGFR levels at genome-wide significance (p<5×10^-8^) were taken forward for genetic association analysis. For the selected genes, a LD-reduced (R^2^ =0.05) set of eQTLs was selected using PLINK v1.90. An inverse variance weighted meta-analysis (IVW) approach was used test the association between predicted gene levels (exposure) and eGFR levels (outcome) using the Mendelian Randomization R package (80). A Bonferroni-corrected association p< (0.05/5 genes=0.01) was considered significant.

### Statistical analysis

Values for **Figure 4, 7, 8** and **S1** were presented as means ± S.E.M. of three independent experiments, data were analyzed by GraphPad Prism (Version 9.3.1) and statistical significance was assessed by an unpaired t test between two means or one-way analysis of variance (ANOVA) was used to assess differences among multiple groups, followed by Tukey’s post hoc test. Results with a p value <0.05 were considered significant.

### Study Approval

The study was approved by the Institutional Review Board at Mass General Brigham and written informed consent was received prior to participation in the study. The trial is registered in ClinicalTrials.gov as NCT 03345446 ‘Circulating RNAs in Acute Congestive Heart Failure (CRUCiAL).

## Data Availability

All data produced in the present study are available upon reasonable request to the authors

## Acknowledgements

This work was funded by grants from AHA (SFRN16SFRN31280008), NHLBI (1R35HL150807-01) and NCATS (UH3 TR002878) to SD.

## Authors’ contribution

EC, RR and VK designed and carried out experiments on the KC with EC leading the study to completion. GPO, GL, PG conducted selected experiments. MS and IL assisted in data analysis related to patient data. MS, TWM and JDM conducted the Mendelian Randomization analysis. QS did the statistical and computational analysis. ICG, JL provided critical review of the manuscript. KK helped design kidney-chip experiments. EC, RR, MS, RS and SD participated in writing the manuscript. RS and SD were responsible for supervision of data analysis and final manuscript. SD was responsible for overall supervision of the experimental design and funding for the project (and is therefore the last author listed).

